# Evaluation of a new dengue 3 controlled human infection model for use in the evaluation of candidate dengue vaccines

**DOI:** 10.1101/2023.06.07.23291100

**Authors:** KK Pierce, SS Whitehead, SA Diehl, G Naro, MC Carmolli, H He, CM Tibery, BP Sabundayo, BD Kirkpatrick, AP Durbin

## Abstract

All four serotypes of dengue virus (DENV) cause the full spectrum of disease. Therefore, vaccines must protect against all serotypes. To evaluate candidate vaccines, a human challenge model of dengue serotype 3 (rDEN30Δ30) was developed. All challenge virus recipients safely met the primary endpoint of viremia and secondary endpoints of rash and seroconversion to DENV-3.

## Background

Infections caused by dengue virus (DENV) are among the most prevalent mosquito borne diseases globally. The World Health Organization (WHO) estimates that dengue viruses cause over 50 million cases of dengue fever (DF) and 500,000 cases of severe disease: dengue hemorrhagic fever or dengue shock syndrome, annually [1]. Effective therapeutic agents for dengue disease do not exist, making vaccine development vital. Disease can be caused by any of four serotypes, DENV1-4, which complicates vaccine development. Infection with one serotype confers long lasting homotypic (serotype-specific) clinical immunity and short-lived heterotypic immunity to the other three serotypes. Although severe disease can occur following primary infection, studies have shown that most severe dengue cases occur in subsequent infections with a serotype different (heterotypic) from the first infection [2]. This phenomenon is thought to be mediated by antibody dependent enhancement of infection (ADE): whereby non-neutralizing antibody from the primary serotype binds but cannot neutralize the second infecting serotype, leading to enhanced cellular entry via the Fcγ receptor and viral replication [3]. For this reason, an ideal dengue vaccine would achieve a homotypic response to all four serotypes.

Several live attenuated tetravalent dengue vaccines are in development. The only licensed vaccine, Dengvaxia®, remains limited in its use due to excess levels of dengue hospitalization following a subsequent dengue infection in individuals who were dengue sero-naïve at vaccination [4, 5].

Given these safety concerns, testing of candidate vaccines in humans must be done with great caution. Human infection models are an established tool for early evaluation of vaccine efficacy. The dengue controlled human infection model (D-CHIM) developed by our team uses well-studied, naturally attenuated dengue strains to elicit safe, low-level dengue infection in healthy volunteers. Our D-CHIM is an infection model with viremia and rash as clinical endpoints rather than a disease model which induces symptomatic illness. To use the D-CHIM for vaccine candidate evaluation, individuals receive a dengue vaccine or placebo and are subsequently inoculated with the challenge strain to determine the protection provided by vaccination. Our first D-CHIM used a recombinant DENV-2 (Tonga) as the challenge strain to assess the protective efficacy of the TV003 vaccine. Six months after vaccination, TV003 elicited complete protection against DENV-2 challenge, whereas all placebo recipients demonstrated infectious DENV-2 (Tonga) viremia following challenge [6].

The success of the DENV-2 D-CHIM compelled us to develop challenge strains for the other DENV serotypes to evaluate the broad protective efficacy of the NIH tetravalent vaccines, TV003 and TV005. We developed a recombinant DENV-3 challenge strain, rDEN3Δ30, for use in D-CHIM studies of candidate dengue vaccines.

DENV-3 Sleman/78 is the parent virus for both the attenuated DENV-3 (rDEN30Δ30/31) vaccine component of TV003 and TV005 and the rDEN3Δ30 challenge strain described herein. DENV-3 Sleman/78 was isolated during a dengue outbreak in 1978 in the Sleman district in Central Java [7]. During that year, illness was mild, mostly observed in children under 15 years of age, with no dengue associated deaths. In contrast, the DENV-3 strain (Bantul), which had circulated the previous year, led to significant illness and hospitalization. The viremia for the Sleman strain was lower than the previous Bantul strain and is considered to be naturally attenuated. The rDEN3Δ30 virus was originally developed as a candidate DENV-3 vaccine. However, in nonhuman primate models, rDEN3Δ30 was not significantly attenuated compared to the parent virus DENV-3 Sleman/78 [8]. rDEN3Δ30 induced viremia in 100% of rhesus macaques with a mean peak titer of 1.5 log_10_PFU/mL compared to 75% of rhesus macaques who were viremic with a mean peak virus titer of 2.0 log_10_PFU/mL following inoculation with the parent virus DENV-3 Sleman/78.

Here we describe the first evaluation of rDEN3Δ30 in healthy, flavivirus-naïve human volunteers and the potential for its use in future D-CHIM studies.

## Methods

This study was conducted under IND application (IND # 16765) at the Center for Immunization Research, Johns Hopkins School of Public Health and the Vaccine Testing Center, University of Vermont College of Medicine, and was approved by their Institutional Review Boards and Biosafety Committees. The National Institute of Allergy and Infectious Diseases (NIAID) Intramural Data Safety Monitoring Board reviewed all study data. Clinicaltrials.gov NCT02684383.

### Study population

Volunteers were recruited from Baltimore, MD, and Burlington, VT. Informed consent was obtained. Similar to previously published studies, inclusion criteria included: healthy subjects age 18-50, negative serology for Hepatitis B, C and HIV, no evidence of prior dengue infection or other flaviviruses, and normal screening values for liver and renal function, urinalysis and coagulation [9]. Women of childbearing potential had a negative pregnancy test at screening and on inoculation day and were required to use a reliable method of contraception for study duration. Following receipt of the test article (rDEN3Δ30 or placebo), subjects recorded their temperature three times daily.

### Study design

A randomized double-blind controlled trial was performed with 14 subjects. Randomization was 5:2, to receive either 3 log_10_ PFU of rDEN3Δ30 or placebo (L-15) as a 0.5 mL subcutaneous injection. To adequately power for viremia only 10 subjects were required to receive DENV-3. Following dosing, volunteers returned to the clinic every other day for the first 2 weeks and then on study days 21, 28, 56, 90, and 180 for clinician assessment, which included physical exam and documentation of clinical signs and symptoms. During the first 2 weeks volunteers were contacted via phone on non-clinic days to review symptoms and to check for fever. During clinic visits, blood was obtained for virus titration and safety laboratory studies (CBC, ALT, coagulation studies). Neutralizing antibody was measured at days 0 (pre-vaccination), 28, 56, 90 and 180 using the 50% plaque reduction neutralization titer (PRNT_50_) assay with DENV-3 Sleman/78 as the target virus [10]. Seroconversion was defined as ≥ 4-fold rise in neutralizing antibody titer at day 90 compared to day 0. Viremia was assessed as described using culture-based method [10]. The frequency of clinical signs and symptoms was compared between rDEN3Δ30 and placebo recipients and their statistical significance was determined using Fisher’s Exact test.

## Results

14 subjects were enrolled, five (35%) were women. At each site five subjects received rDEN3Δ30 and two subjects received placebo (diluent). Mean age was 34 years (range 24-47 years).

All 10 subjects who received rDEN3Δ30 (100%) met the primary endpoint of viremia and 8 (80%) had a dengue-like rash. Rash was only seen in rDEN3Δ30 recipients and was dengue-like in appearance (diffuse, maculopapular, and present over the trunk and extremities). It was observed approximately four days after challenge, and completely resolved within 7-10 days. Among the eight rash presentations, five were graded as moderate and three were mild. Transient, asymptomatic neutropenia and thrombocytopenia were seen in one and two volunteers, respectively.

Individual viremia data are presented in Figure 1A. The mean peak viremia titer in rDEN3Δ30 recipients was 1.8 log_10_ PFU/mL (range 0.5-2.1); mean day of onset was 4.6 days (range 3-6 days) with a mean duration of 3.4 days. All 10 rDEN3Δ30 recipients seroconverted to DENV-3; the PRNT_50_ to DENV-3 peaked at day 28. Although titers declined slightly, they remained high at day 180 with a GMT of 143 (range 44-392). Peak low level heterotypic antibodies to serotypes DENV-1 and -2 were seen in rDEN3Δ30 recipients **(Figure 1B)**.

**Figure 1.**
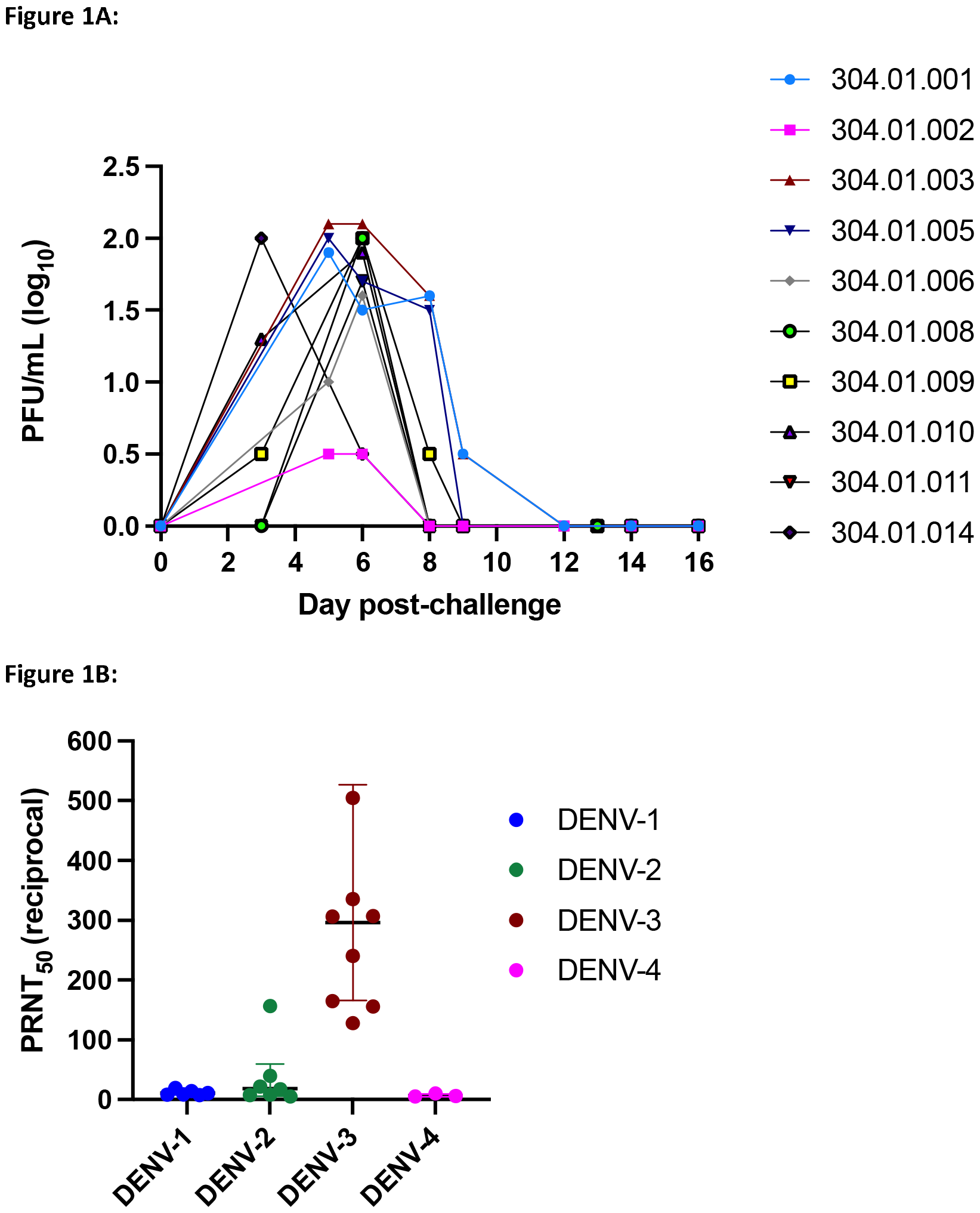
**(A)** rDEN3Δ30 was cultured from serum samples collected on the indicated days. Virus was detected by culture in Vero cells as described in the methods. Time course of viremia for the 10 individuals administered rDEN3Δ30 is shown. The lower limit of detection is 0.5 log_10_ PFU/mL of serum. PFU, plaque forming unit. **(B)** Serum was collected from the 10 infected individuals on study days 0, 28, 56, 90, and 180. The peak PRNT_50_ achieved through study day 90 for each of the four DENV serotypes is shown for all individuals who developed a PRNT_50_ > 1:5 (limit of detection). The geometric mean peak titer with SD is indicated.

Overall, the DENV-3 inoculation was well-tolerated. Fever was not observed in any volunteer. Local reactogenicity in the rDEN3Δ30 group included injection site erythema (10%) and injection site induration (20%). The frequency of clinical signs and symptoms other than rash were not significantly different between the groups (**Table 1**). Of the 86 reported events, 77 (89.5%) were considered mild; the remainder were graded as moderate. The only moderate events deemed to be related to rDEN3Δ30 or placebo were rash and fatigue.

**Table 1:**
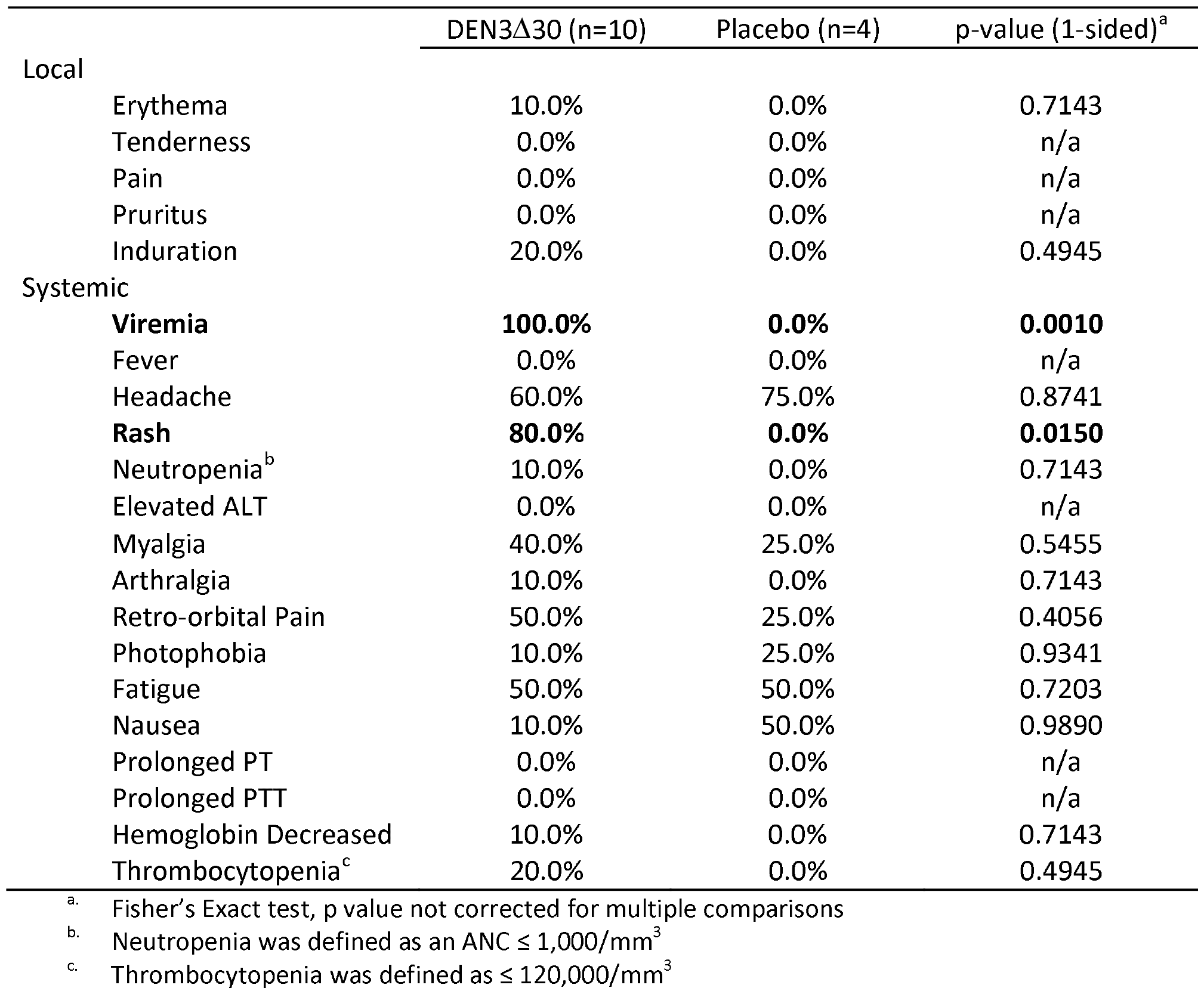
Clinical signs and symptoms following a single dose of rDEN3Δ30 compared with placebo.

## Discussion

Important requirements for development of a safe dengue vaccine include the need to achieve a robust and balanced immune response protective against all four serotypes while maintaining an acceptable reactogenicity profile. The implication of an incomplete or imbalanced response, as recently seen in long-term follow up of Dengvaxia® recipients, is the possibility the vaccine may predispose to more severe disease upon subsequent exposure to DENV years following vaccination [4]. Although neutralizing antibody is thought to be a mediator of protection, titers of neutralizing antibody induced by Dengvaxia® were not indicative of protection against dengue [11]. Use of the D-CHIM to evaluate the protective efficacy of candidate vaccines offers additional assurances of serotype-specific protection in advance of phase III testing in large endemic-setting populations.

Our study demonstrated rDEN3Δ30 is highly infectious and immunogenic yet induces mild to moderate clinical signs in all inoculated volunteers. The infectivity and safety profile of the DENV-3 challenge virus makes it ideal for use in CHIM studies to evaluate candidate vaccines. As seen in the DENV-2 CHIM, we targeted induction of detectable viremia in a high percentage of subjects, with peak titers that elicited only mild to moderate clinical signs [6]. Prior to our model, another research team performed a Dengue 3 CHIM as a disease model which caused symptomatic dengue-like illness. Volunteers in that study experienced fever for several days, transaminitis, and one subject had evidence of perihepatic and pericardial effusions by ultrasound [12]. In our study, 100% of rDEN3Δ30 recipients had detectable viremia and seroconverted to DENV-3 based on neutralizing antibody; dengue-like rash was seen in eight of 10 volunteers (80%). Neither fever, transaminitis, nor effusions were observed in any volunteer.

Given that 75% of DENV infections do not present for clinical care, we propose that the endpoints of viremia and rash are relevant to the course of natural infection and are reproducible targets upon which candidate vaccines can be safely evaluated.

When compared to our DENV-2 D-CHIM, rDEN3Δ30 infection induced a lower mean peak virus titer (1.77 log_10_ PFU/mL) vs. rDEN2Δ30 (2.3 log_10_ PFU/mL). Nevertheless, rDEN3Δ30 induced rashes of moderate severity in 50% of challenge volunteers. This possible disconnect of peak viremia titer and rash severity will be monitored closing in subsequent trials.

The rDEN3Δ30 CHIM is an important new tool for evaluating candidate dengue vaccines in advance of phase III efficacy trials in endemic populations. Its safety and infectivity profiles are acceptable and because of rDEN3Δ30’s high infectivity, efficacy studies can be powered for significance with relatively few subjects. We argue that, in light of the concerns raised with the most recent large scale Phase III dengue vaccine trials, the CHIM provides a platform to test vaccine safety and efficacy on a smaller scale before large scale vaccine roll out. This DENV-3 CHIM is currently being used to evaluate the efficacy of the TV005 vaccine to protect against infection with rDEN3Δ30 at 6 months post-vaccination.

## Data Availability

All data produced in the present work are contained in the manuscript

## Acknowledgments

Dengue virus strains were provided by the Laboratory of Infectious Diseases, NIH. We would like to thank all of the dedicated volunteers, and the excellent research nurses and staff at the JHU CIR and UVM VTC Clinical Research Centers. In addition, the authors would like to thank the Office of Clinical Research Policy and Regulatory Operations at the NIH for providing the regulatory oversight.

